# Projected ecological and disruptive impacts of climate change on malaria in Africa

**DOI:** 10.1101/2025.02.11.25322113

**Authors:** Tasmin L. Symons, Alexander Moran, Ann Balzarolo, Camilo Vargas, Mairi Robertson, Jailos Lubinda, Adam Saddler, Michael McPhail, Joseph Harris, Jennifer Rozier, Annie Browne, Punam Amratia, Amelia Bertozzi-Villa, Samir Bhatt, Ewan Cameron, Nick Golding, David L. Smith, Abdisalan M. Noor, Susan F. Rumisha, Matthew D. Palmer, Daniel J. Weiss, Naomi Desai, David Potere, Nicholas Sukitsch, Wendy Woods, Peter W. Gething

**Affiliations:** School of Population Health, Curtin University, Bentley, Western Australia; Malaria Atlas Project, The Kids Research Institute Australia, Nedlands, Western Australia; Boston Consulting Group; Malaria Atlas Project, Ifakara Health Institute, Dar es Salaam, Tanzania; Institute for Disease Modelling, Gates Foundation, Seattle WA, USA; Department of Infectious Disease Epidemiology, Imperial College London, UK; University of Copenhagen, Denmark; School of Physics, Mathematics and Computing, University of Western Australia, Western Australia; Institute for Health Metrics and Evaluation, University of Washington, Seattle WA, USA; Harvard T.H. Chan School of Public Health, Boston, MA; Applied Health Analysis for Delivery and Innovation (AHADI), Boston, MA; Met Office Hadley Centre, Exeter, UK; School of Earth Sciences, University of Bristol, Bristol, UK

## Abstract

The implications of climate change for malaria eradication in the 21^st^ century remain poorly resolved. Many studies have focussed on parasite and vector ecology in isolation, neglecting the interactions between climate, malaria control, and the socioeconomic environment, including the disruptive impact of extreme weather. Here we integrate 25 years of data on climate, malaria burden, control interventions, socioeconomic factors, and extreme weather events in Africa. Using a geotemporal model linked to an ensemble of climate projections under the Shared Socioeconomic Pathway 2-4.5 (SSP 2-4.5) scenario, we estimate the future impact of climate change on malaria burden in Africa, accounting for both ecological and disruptive effects. Our findings suggest climate change could lead to 123 million (projection range 49.5 million - 203 million) additional malaria cases and 532,000 (195,000 - 912,000) additional deaths in Africa between 2024 and 2050 under current control levels. Contrary to the prevailing focus on ecological mechanisms, extreme weather events emerge as the primary driver of increased risk, accounting for 79% (50-94%) of additional cases and 93% (70%-100%) of additional deaths. Most increases are due to intensification in existing endemic areas rather than range expansion, with significant regional variation in impact. These results highlight the urgent need for climate-resilient malaria control strategies and robust emergency response systems to safeguard progress toward malaria eradication in Africa.

## Main text

In the 21^st^ century, 15 years of declining malaria burden in Africa have been followed by a decade of faltering progress. Growing optimism around new vector control, chemoprevention, and vaccine interventions – along with increasingly sophisticated surveillance and deployment capabilities – is being tempered by the concurrent challenges of uncertain financing, rapidly growing populations at risk, and intensifying biological threats. At this critical juncture, a question of profound importance is the extent to which climate change threatens efforts to regain momentum against the disease and the feasibility of eradication as a mid-century goal.

The link between climate and malaria is widely accepted and extensive research has elucidated different causal pathways and sought to project future impacts^1,2^. Such research has permeated local and global malaria policy formulation^3,4,5,6^, but the ongoing lack of consensus on the directionality, magnitude, and geographical distribution of climate change effects hinders the formulation of detailed response strategies ^2,7^.

Climate mediates malaria vector and parasite ecology through multiple mechanisms. A long lineage of studies have used entomological observations and mathematical models to explore the effects of ambient temperature on *Anopheles* mosquito lifespan, blood meal frequency and other life history characteristics^8,9^, the duration of *Plasmodium* extrinsic incubation period^10^, and the combined implications for vectorial capacity and malaria transmission intensity^8,9,10,11,12,13,14^. Recent advances have tailored this understanding to the most important malaria vectors in Africa – members of the *Anopheles gambiae* complex^15^ - and the invasive vector *Anopheles stephensi*^16,17^. Other work has focused on understanding the way climate affects the abundance of emergent adult mosquitoes by influencing the availability of mosquito larval habitat for oviposition, and the suitability of that habitat to sustain development of the larval stages^18,19,20,21^.

### Climate-malaria impacts must be explored in context

As understanding of climate effects on malaria transmission ecology has developed, many studies have extrapolated those mechanisms under future climate scenarios to predict how malaria may respond to climate change. Some studies have incorporated temperature effects only^22,23^, while others have used more comprehensive empirical or mechanistic models to capture temperature, rainfall, and humidity effects on both larval and adult mosquito stages^20,24,25,26,27,28,29^. Most commonly, these efforts have aimed to predict changes in the geographical (or seasonal) boundaries of transmission^20,22,24,27^. However, many areas of the world are already suitable for malaria transmission yet have none whilst, within existing boundaries, transmission varies by several orders of magnitude. Understanding boundary changes therefore provides only partial insight into climate change impacts on malaria.

Other studies have sought to use mechanistic models to predict climate change impacts on transmission in absolute terms, based either on extrapolated parametrisations from laboratory or field observations^25,26,27,30^, or calibration to local observations of entomological inoculation rate, asexual blood-stage infection prevalence, or case incidence^28,29,31^. However, such models show limited ability to reproduce historical endemicity^32^ or observed patterns of malaria infection prevalence across Africa, casting doubt on their reliability for projecting future changes in those patterns^20,26,33^.

Nearly all existing projections share a central limitation: while they explore climate effects in isolation, they do not adequately account for non-climate determinants of malaria trends. Before the roll-out of modern antimalarial medicines, rising drug resistance in the 1990s contributed to increasing malaria trends in many locations, including those intensively studied for climate change effects^34,35^. Subsequently, aggressive scale up of effective interventions between 2000-2015 almost halved mean infection prevalence across the continent, albeit unevenly^36^. In recent decades malaria-endemic Africa has rapidly urbanised and developed, with over 40% of the population now living in cities^37^, and more than a quarter now benefiting from improved housing^38^. These factors have fundamentally reshaped the modern-day landscape of malaria risk, driven long-term changes independently of climate trends, and mean that identical climate conditions can host very different levels of malaria transmission^39,40^. Failing to account for these factors prevents an accurate assessment of the role of climate — and, by extension, climate change — in shaping future malaria risk.

Importantly, the built environment and malaria control efforts not only mediate transmission independently of climate but also provide additional pathways by which weather and climate can impact malaria. Extreme weather events like floods and cyclones damage homes and infrastructure, disrupting access to healthcare, protective housing, and malaria control. Malaria surges linked to recent extreme weather events in Africa and Asia have been widely documented^41,42^, including causal studies on the impact of disrupted malaria control, even after substantial emergency interventions^41^. In Africa, climate change is predicted to lead to more frequent and severe floods, and more severe southern Indian Ocean cyclones^43^. With previous climate-malaria work focussing almost exclusively on ecological mechanisms, no study has attempted to quantify the disruptive impact that intensifying extreme weather events might have on malaria and its control in Africa.

### A framework for projecting the impact of climate change on malaria

Here we provide the first projections of possible impact from both ecological and disruptive effects of climate change on malaria in Africa. Crucially, we first characterise the climate-malaria relationship in the context of the other key determinants of malaria risk, based on 25 years of comprehensive data on malaria infection prevalence, intervention coverage and socioeconomic conditions. A schematic overview of our analytical framework is shown in Extended Data Fig. 1. Bias-corrected and downscaled Coupled Model Intercomparison Project Phase 6 (CMIP6) member global climate model (GCM) outputs obtained from the NASA Earth Exchange Global Daily Downscaled Projections provided future projections of climatic variables to 2050^44^, and were calibrated to observed climate data^45,46,47^ from the co-observed 2000-2014 period, generating consistent timeseries of climatic variables from 2000-2050 at monthly timesteps and 5×5 km spatial resolution. Between-GCM uncertainty and variability were accounted for using an ensemble of CMIP6 members – fourteen models for evaluating ecological impacts, and a tractable subset of three for disruptive impacts. We focussed analysis on the ‘middle of the road’ scenario SSP 2-4.5, designed to be broadly consistent with current international pledges on reduced greenhouse gas emissions^48^.

Ecologically-driven malaria impacts were assessed by first using mechanistic models to transform the processed climate data into two indices representing the climate-malaria interface: (i) the effects of temperature on mosquito and parasite lifecycles and their interaction; and (ii) the interaction of rainfall, humidity and temperature to determine the relative availability of transient larval habitat for mosquito oviposition and larval development. Use of monthly timesteps enabled appropriate propagation of changing regimes of seasonal, inter-annual, and inter-decadal climate variation. Second, for the period of observational record in the training data (2000-2022), the two climatic indices were used as geotemporal predictor variables, along with gridded data on mosquito relative species abundance^49^, permanent larval habitat availability, and geotemporal estimates of vector control coverage^50^, seasonal malaria chemoprevention (SMC), antimalarial drug treatment^51^ and improved housing^38^ in a hierarchical Bayesian geotemporal model fitted to 49,994 geo-located observations of malaria infection prevalence (*Plasmodium falciparum* parasite rate: age- and diagnostic-standardised parasitaemia rates in 2-10 year olds, henceforth *Pf*PR_2-10_) collated across Africa by the Malaria Atlas Project^51^. This framework estimated empirically the relative and absolute effects of each predictor variable, including their interactions, on malaria transmission. Third, holding malaria control coverage and socioeconomic metrics at present-day levels, the fitted model was used to generate both SSP 2-4.5 and counterfactual (i.e. projecting present-day conditions with no future climate change) *Pf*PR_2-10_ scenarios for each climate ensemble member from 2024 to 2050, which were then differenced to generate estimates of climate change impact (See detailed scenario definitions in Extended Data Table 1).

Disruption-driven malaria impacts were assessed by first simulating plausible scenarios of flooding and cyclone events in sub-Saharan Africa consistent with both past observations^52,53^ and GCM-projected future climate using statistical models calibrated and validated to historical events. For fluvial and pluvial flooding, the frequency, duration and extent of observed flood events were associated with a range of climate variables to generate a set of geotemporal catalogues of simulated future flood events under both counterfactual (i.e. without future climate change) and SSP 2-4.5 conditions to 2050. Similarly, the statistical relationship between climate variables and past Indian Ocean cyclone intensity and track morphology was characterised using methodology and datasets from a recently updated storms model^54^ which was also then projected to generate SSP 2-4.5 and counterfactual event catalogues to 2050.

The impact of individual weather events on malaria has almost never been measured directly – in part because of associated disruptions to care seeking and disease surveillance systems. Instead, we assembled both qualitative and quantitative records of past extreme weather events in malaria endemic settings that detailed the type and magnitude of damage and disruption experienced, and supplemented these with insights from 35 stakeholder interviews, capturing first-hand accounts of malaria impacts on the ground, including from representatives of humanitarian response agencies, national malaria programs, and global health agencies. This mixed-methods approach identified four primary pathways by which extreme weather events can impact malaria: via damage or disruption to the protective effects of (i) improved housing, (ii) indoor vector control tools, or (iii) access to antimalarial treatment; or (iv) from changes to *Anopheles* larval habitat availability due to flood water (Extended Data Table 2). For i-iii, we used the compiled evidence (Extended Data Table 3) to define plausible levels of disruption by event type, duration and intensity - distinguishing between acute disruption in the immediate aftermath, and a period of persisting disruption before return to pre-event conditions (see Extended Data Fig. 2 for a schematic overview of our approach). To reflect the inherent variability and uncertainty in these disruption parameters, we also generated ranges spanning 50% to 150% of the central consensus values. Combining these elements, we generated modified versions of the geotemporal data layers for vector control coverage, antimalarial drug treatment, and improved housing that incorporated disruptions under both SSP 2-4.5 and counterfactual scenarios to 2050. Larval habitat impacts contemporaneous with extreme events (iv) were captured via the habitat suitability model described earlier. The fitted hierarchical Bayesian geotemporal model was then used to reproject *Pf*PR*_2-10_* with these disruptive impacts, both separately and in combination with the ecological impacts, which were then differenced from counterfactual scenarios without future climate change (Extended Data Table 1). Uncertainty in the damage parameter ranges was combined with GCM model ensemble spread to derive overall projection ranges (Extended Data Fig. 6). All projections of *Pf*PR_2-10_ were converted into estimates of malaria clinical incidence using an established natural history model^55^ and with gridded future population projections consistent with SSP 2-4.5^56^. Mortality projections were derived from projected untreated cases and WHO estimates of untreated case-fatality rates^57^.

### Ecological and disruptive impacts of climate change on malaria transmission

We project that, considered in isolation, the ecologically-driven impacts of climate change on malaria transmission would lead to minimal overall change in Africa by 2050 under SSP 2-4.5, with a continent-wide population-weighted mean percentage point increase in *Pf*PR_2-10_ due to ecological impacts of just 0.12% relative to the counterfactual without future climate change (14-model ensemble spread −0.03% to 0.33%). However, this continental aggregation masks extensive geographical variation (Fig. 1a-c), with much larger local and regional impacts predicted. We project that warming under SSP 2-4.5 would increase malaria risk in regions where lower temperatures currently constrain transmission: the belt of lower latitude Southern Africa including Angola, Zambia and southern Democratic Republic of Congo (DRC); and highland areas in Ethiopia, Kenya, Rwanda, Burundi and eastern DRC (Fig. 1c). Conversely, warming would drive reduced transmission in Sahelian regions including in Burkina Faso, Niger, northern Nigeria, and Chad as temperature regimes would increasingly exceed optimal ranges for *Anopheles* survival (Fig. 1b). Changing temperature suitability (Fig. 1d) rather than larval habitat availability (Fig. 1e) was identified as the dominant ecological mechanism, although this was partly driven by the greater GCM ensemble divergence for rainfall projections versus those for temperature: individual GCM projections did include more substantial habitat impact (Extended Data Fig. 3).

**Fig. 1:**
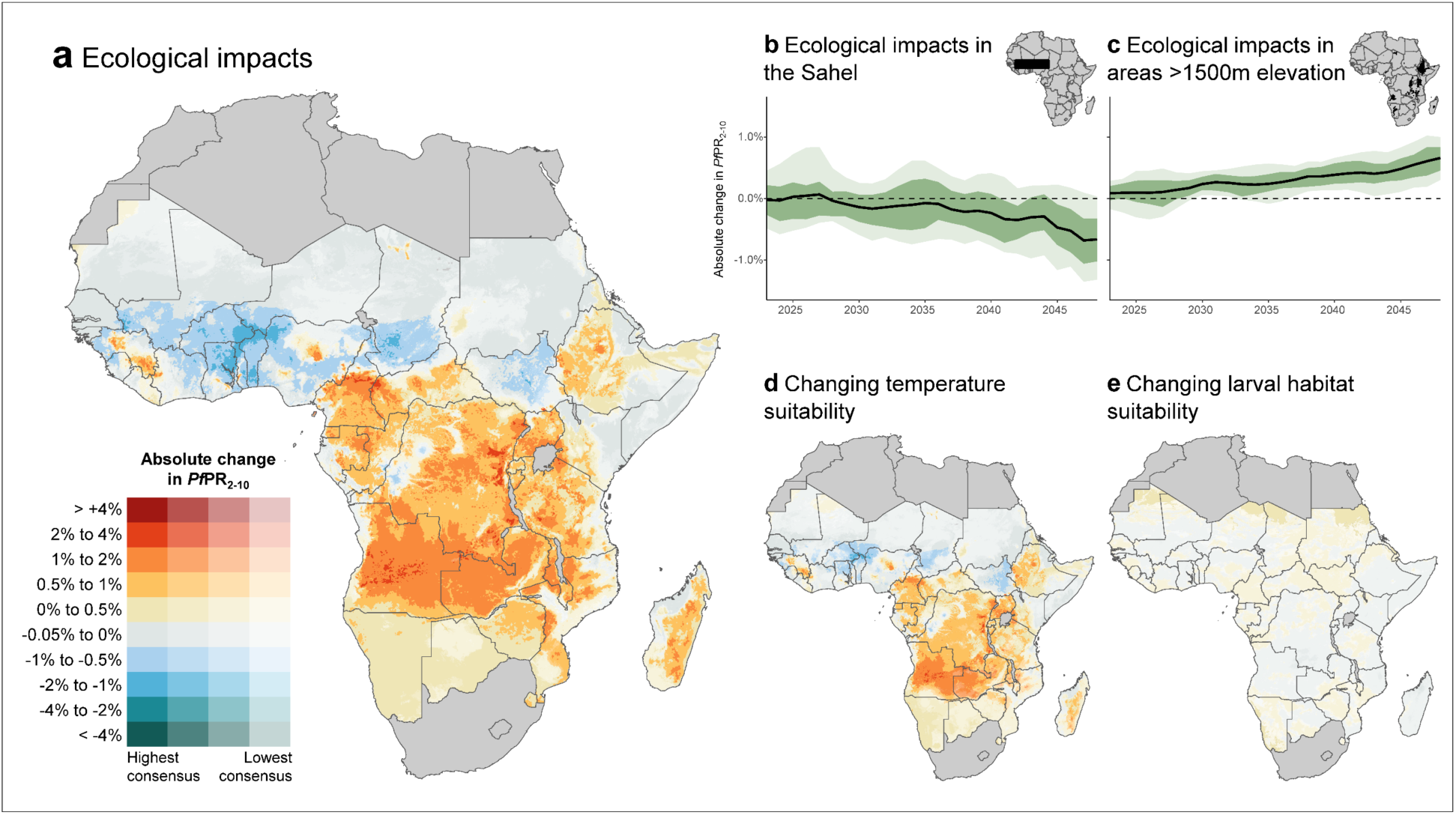
Projected ecologically-driven impacts of climate change on malaria transmission under SSP 2-4.5. **a**, Mapped percentage point change in *Pf*PR_2-10_ due to ecologically-driven impacts of climate change. Values compare annual median across 2040-2049 under SSP 2-4.5 scenario (ensemble mean of 14 downscaled and bias-corrected CMIP6 members) to counterfactual (no future climate change) scenario, with intervention coverage and socioeconomics held constant at present-day levels in both scenarios. Large water bodies and out-of-scope countries are masked in dark grey. **b**,**c**, Time-series of population-weighted 3-year rolling mean change in *Pf*PR_2-10_ from 2019-2022 baseline in the Sahel (**b**) and areas of mean elevation above 1500m (**c**), with ribbons representing ensemble 25^th^-75^th^ (darker green), and 10^th^-90^th^ percentile (lighter green) and inset maps showing geographical subsets. **d**,**e**, Decomposition of ecological impacts due to changing temperature suitability (**d**) and changing larval habitat suitability (**e**), with colour ramping identical to panel (**a**). Individual ensemble member summaries are shown in Extended Data Fig. 3.

Our simulated flood event catalogues reflected a net increase in flooding across Africa due to climate change, with area-days flooded 13% (ensemble spread 9% - 17%) greater in the 2040s under SSP 2-4.5 versus present-day conditions (Extended Data Fig. 4). We projected Indian Ocean cyclones to shift in severity under SSP 2-4.5 versus present-day conditions– with fewer Category 1-4 events but more frequent Category 5 (Extended Data Fig. 4). Although there is substantial uncertainty reflected in the ensemble spread, our projections are consistent with current consensus and reflect expectations of increased atmospheric moisture transports and intensification of the water cycle^6,43^, although we note that evidence on changing extremes of precipitation is considered stronger than that for Indian Ocean cyclone systems. We project that, under SSP 2-4.5, the disruptive impact of intensifying extreme weather events in the 2040s could, without mitigation measures, lead to *Pf*PR_2-10_ increasing across 24% of the land area of malaria-endemic Africa, associated mainly with major river systems and the cyclone-prone coastal regions of southeast Africa (Fig. 2a).

**Fig. 2:**
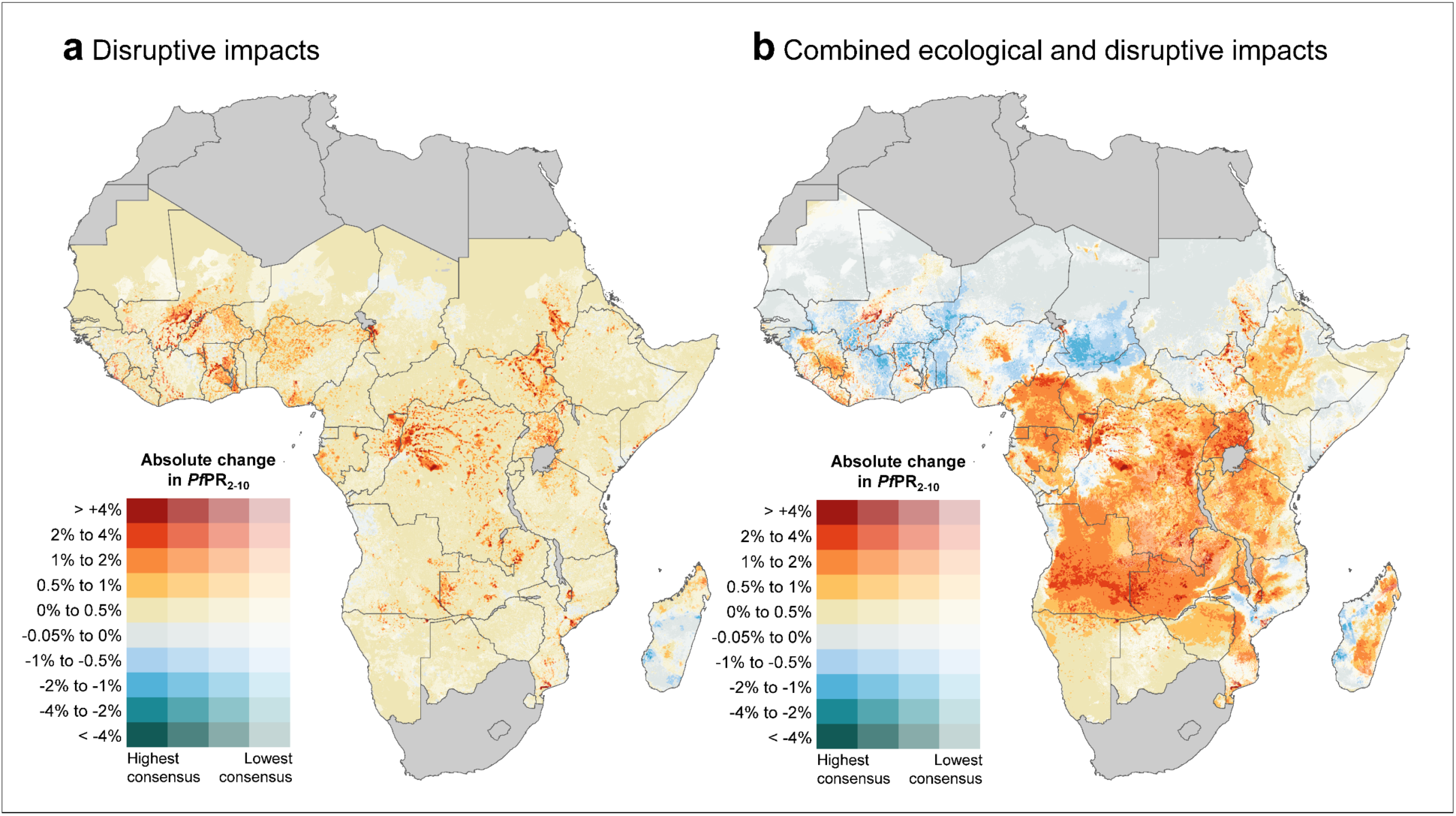
Projected disruptive and combined disruptive + ecological impacts of climate change on malaria transmission in the 2040s. **a**, Mapped percentage point change in *Pf*PR_2-10_ due to disruptive impacts of climate change only. Values compare *Pf*PR_2-10_ median across 2040-2049 impacted by simulated extreme weather events consistent with the SSP 2-4.5 scenario (ensemble mean of three downscaled and bias-corrected CMIP6 members) versus a counterfactual scenario with extreme weather events projected at present-day levels. Intervention coverage and socioeconomics otherwise held constant at present-day levels. Large water bodies and out-of-scope countries masked in grey. **b**, Mapped percentage point change in *Pf*PR_2-10_ due to combined disruptive and ecological impacts of climate change. Values compare *Pf*PR_2-10_ median across 2040-2049 under the SSP 2-4.5 scenario versus the counterfactual (no future climate change) scenario.

The combined ecological and disruptive impacts of climate change on *Pf*PR_2-10_ projected to the 2040s are shown in Fig. 2b, and Extended Data Fig. 5. In several settings – notably Uganda, eastern DRC, Zambia, and Angola – the ecological and disruptive effects compound, with ecologically-driven increases in underlying transmission exacerbated by more frequent extreme weather events that further expose those vulnerable populations. Across floodplains in the Sahel, meanwhile, projected decreases in ecological suitability are negated by more frequent and severe flooding, for example along the Niger River in Mali and Nile in South Sudan. In combination, we project that ecological and disruptive impacts in the 2040s under SSP 2-4.5 would lead to increased malaria transmission for 67% of Africa’s population, with around 73M exposed to a percentage point increase in *Pf*PR_2-10_ in excess of 2% (Extended Data Fig. 5).

### Increased malaria burden due to climate change

Over the next twenty-five years, we project that climate change - through combined ecological and disruptive impacts - could lead to 123 million additional clinical cases of malaria in Africa (projection range 49.5 million-203 million) under SSP 2-4.5 (Fig. 3a). By the 2040s, this equates to an increase of 0.4-2.6% in case incidence rate, relative to the counterfactual without future climate change. The growing disruptive impact of more frequent and severe extreme weather events contributes 79% (projection range 50-94%) of this projected rise in case incidence - more than three times greater than the contribution of ecologically-driven impacts (21%, 6-50%). Of the disruption-driven impacts to case incidence, those resulting from reduced access to malaria treatment form the largest component (37.8% of total, ensemble spread 34.2-41.3%), followed by damage to housing (23.4%, ensemble spread 21.3 - 27.1%) and disruptions to vector control (14.9%, ensemble spread 14.6-15.5%) (Fig. 3a). Fig. 3b maps the cumulative change in clinical cases by 2049 for each 5×5 km grid cell. Some locations with large projected rises in transmission rates occur in relatively sparsely populated regions – for example central Angola and western Zambia – and hence incur only modest increases in cases. Elsewhere, substantial climate impact coincides with regions of dense populations at risk, most notably southern and central Nigeria, and the African Great Lakes region encompassing parts of Kenya, Uganda, eastern DRC, Rwanda and Burundi. Assuming present-day untreated case fatality ratios^57^, our projected changes to case incidence and treatment rates would equate to 532,000 (195,000 - 912,000) additional malaria deaths over the same period, a relative increase in annual mortality rates of 0.9 - 5.4% compared to the counterfactual in the 2040s. As would be expected, disruptions to case management account for an even greater proportion of this increase in deaths (65%, ensemble spread 62%-70%) than for case incidence.

**Fig. 3:**
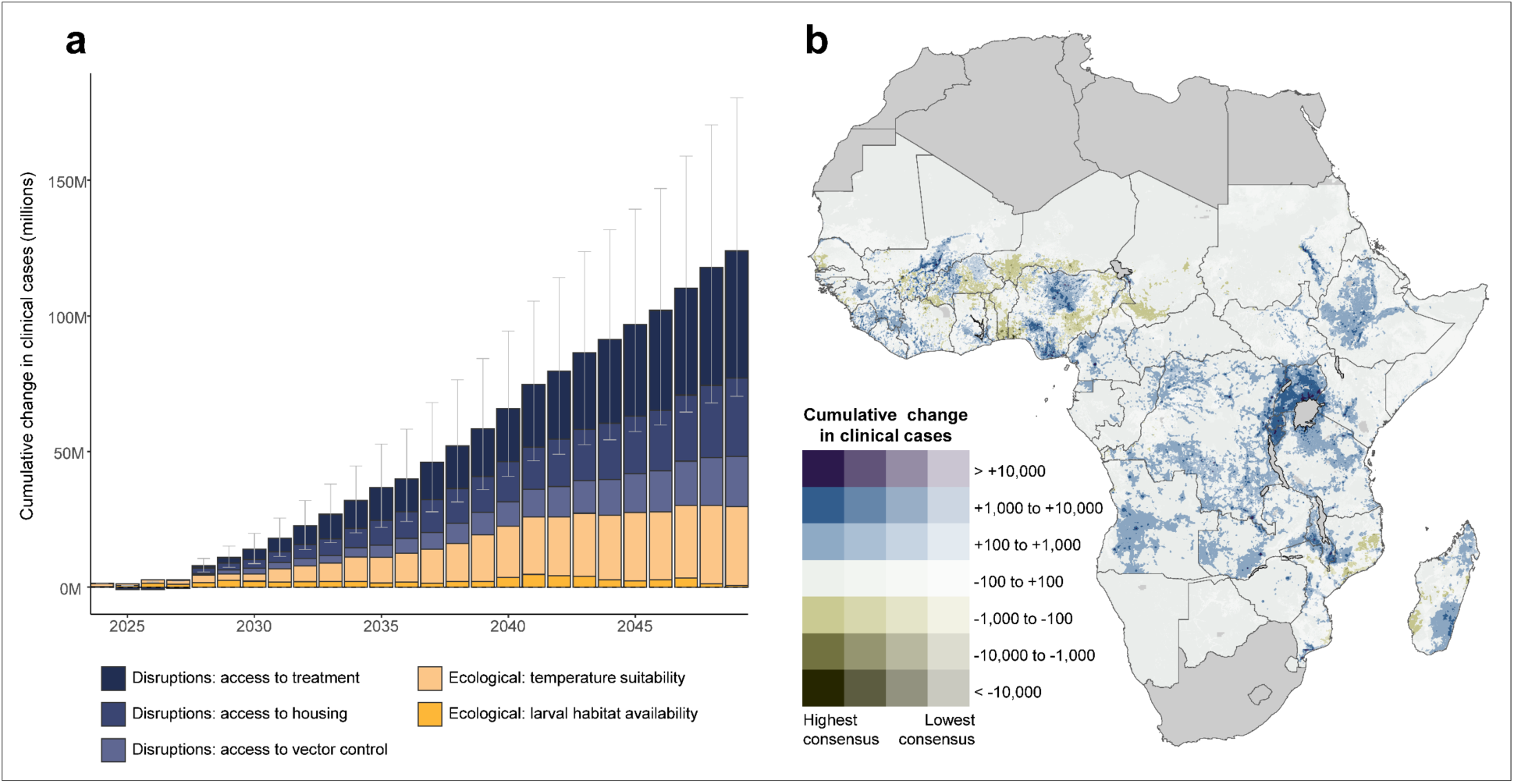
Projected cumulative impact on clinical malaria incidence 2024-2049 from climate change under SSP 2-4.5. **a**, Projected continent-wide cumulative impact on cases by year, with decomposition showing distinct contributions of the different ecological and disruptive drivers of climate change impact. Grey bars denote projection range, where this is calculated as the 10^th^ and 90^th^ percentile of the model ensemble across all GCMs and disruption parameter ranges. **b**, Mapped cumulative impact on cases with colour scale additionally reflecting degree of GCM ensemble consensus. Large water bodies, national parks with extremely low or zero population, and out-of-scope countries are masked in dark grey. All cumulative impacts are relative to 2022 case counts, aligned to national totals reported in the 2023 World Malaria Report^57^.

Many preceding studies have focused on identifying climate change-driven changes to the boundaries of transmission suitability. However, we project that the great majority of additional cases would occur in settings already suitable for transmission, with only 0.05% (33,000 cases, three GCM ensemble spread 20,000-51,000, 0.03-0.07%) of additional cases in the 2040s occurring in places currently outside suitability limits.

## Discussion

This analysis has, for the first time, examined the potential threats posed by ecological and disruptive impacts of climate change on malaria transmission and burden in Africa, quantified the plausible magnitude of these impacts in absolute terms, and done so while placing climate mechanisms in the context of other key factors mediating malaria risk. While the projected impacts on transmission are ostensibly modest, we have shown that, when translated into impact on clinical incidence and deaths over the coming decades, the implications of climate change become substantial, potentially resulting in hundreds of millions of additional cases and hundreds of thousands of additional deaths over the next 25 years. In the wider context of plans for accelerated burden reduction and eradication over that timescale, such climate impacts would be significant and demand multisectoral policy responses.

In our analysis of ecologically-driven climate impacts, we have taken current state-of-the-art understanding of the main biophysical mechanisms involved and, uniquely, used detailed historical malaria, intervention, and broader contextual data to estimate the absolute contribution of these mechanisms to realised malaria transmission. We have also demonstrated that a narrow focus on changing transmission boundaries ignores around 99% of likely climate impact in Africa, which occurs within regions already experiencing transmission.

Unlike the analysis of ecological impacts, our modelling of disruption-driven impacts had no precedent in the literature to build upon or compare against. The comparative paucity of observational data and understanding of extreme weather effects on malaria necessitated a more heuristic approach to quantifying likely impacts. Nonetheless, by compiling quantitative and qualitative evidence including the first-hand accounts of those witnessing and responding to extreme weather events in Africa, we have identified mechanisms and plausible magnitudes of impact that likely reflect real-world experience. By combining these effects with simulated future events consistent with GCM projections, we have provided the first systematic exploration of the impact on malaria of worsening extreme weather associated with climate change. Importantly, we identify these disruptive impacts to be around three times larger than ecologically-driven effects.

This analysis should be interpreted in the context of its design, assumptions, and limitations. (i) Our results reflect multi-model ensemble means across decadal timespans. This choice reflects a balance between reducing artefactual inter-model and inter-annual variation while addressing the medium-term timescales relevant to urgent malaria policy needs. Decadal summaries are, by definition, also subject to inter-decadal climate variability known to influence malaria^58^, but our ensemble approach minimises their artefactual impacts. (ii) We opted to use a four-year period (2019-2022) as our present-day climate baseline. Again, this choice sought to balance the need to reduce potential artefacts introduced by short term or stochastic year-on-year variation in our chosen reference period versus the imperative to make comparisons of future malaria landscapes against present-day conditions. (iii) We limited our analysis of disruptive impacts to floods and cyclones, for which the evidence base of impact on transmission is best-established^41,42^. Future climate change will conceivably impact malaria via a wide range of indirect mechanisms such as food insecurity, loss of livelihoods, conflict, health system stressors and economic disruption. While such mechanisms have not been included in this analysis, it is important to recognise their potential significance alongside the more tractable mechanisms we were able to address. (iv) Besides omission of these broader factors, we also deliberately hold constant present-day levels of transport and healthcare infrastructure, housing quality and malaria control – accounting for direct climate impacts on these determinants but not background trends. As such our projections allow exploration of climate change effects but are not intended as forecasts of future conditions.

Taken as a whole, our findings demonstrate the potential of climate change to substantially hinder malaria reduction and eradication over the coming 25 years, while emphasizing that the ultimate outcome will hinge on the effectiveness of malaria control and the resilience of the health systems delivering it. While the projected scale of impacts from gradually changing transmission ecology could likely be counteracted with modest additional intervention, increasing damage and disruption to control efforts from extreme weather poses a more profound threat. Mitigating this threat will require renewed focus on climate-resilient control strategies at international and local levels, encompassing both reinforced emergency early warning and response mechanisms as well as locally tailored use of new tools less vulnerable to climate disruption^59^.

Eradicating malaria in the first half of this century would be one of the greatest accomplishments in human history. Accelerating climate change poses multiple threats to this ambition. In this context, climate-proofed eradication strategies will demand ongoing vigilance, proactive planning, engaged communities, and sustained financing, all within the broader framework of robust health systems and climate-resilient societies.

## Detailed methods

Our analysis framework comprised nine main stages, summarised in Extended Data Fig. 1.

### 1. Preparation of consistent geotemporal climatologies, 2000-2050

#### Historical climate data

Climate data were obtained from the Climate Hazards Centre^45^ and Climate Research Unit gridded Time Series^46^, downscaled by Worldclim^47^. Data were gap-filled^60^, aligned to a standard 5×5 km reference grid, and aggregated to monthly time-steps. Details of all historical climate data used in this study are provided in Supplementary Annex Table 1.

#### CMIP6 projections

Projections of future climate under SSP 2-4.5 were obtained from the NASA Earth Exchange Global Daily Downscaled Projections (NEX-GDDP-CMIP6)^44^. These data consist of downscaled and bias-corrected daily CMIP6 multi-model ensemble outputs for historical (2000-2014) and future projection (2015-2050) eras. Data were aggregated to monthly resolution before applying the delta method^61^ to provide a final calibration to historical climate data described above. To account for between-model uncertainty, fourteen models were processed and used for projection of ecologically-driven climate impacts (see Supplementary Annex Table 2), while a thinned subset of three models (ACCESS-CM2, EC-Earth3-Veg-LR and MPI-ESM1-2-LR) was used for the additional analysis of disruptive climate impacts.

### 2. Modelling of historical and projected climate suitability indices

The assembled climate data were used in two mathematical models to develop two geotemporal suitability indices: (i) a temperature suitability index (TSI) tracking relative vectorial capacity, and (ii) a larval habitat suitability index (HSI) measuring relative availability and potential productivity of mosquito larval breeding sites.

Temperature provides both an upper and lower constraint on malaria transmission, reflecting the ectothermic nature of mosquito and parasite lifecycles. Mathematical models linking temperature to relative vectorial capacity are well-established and described elsewhere^62^. Here we updated a degree-day-based framework^63,64^ to include recently published data on the *Anopheles gambiae* complex^15^. The resulting temperature suitability curve is nonlinear, peaking at 26.4°C.

To describe the relationship between local rainfall, temperature and humidity and the resulting availability of habitat for oviposition and larval development we discretised a Clausius-Clapeyron-based model of habitat availability used in an established mechanistic model of malaria^65^ as follows: let *R*_*t*_ denote rainfall volume at time *t*, so that transient larval habitat is given by the recursion:

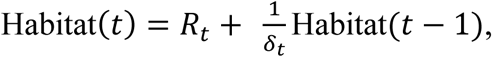

where *δ*_*t*_ is the time-dependent temperature- and humidity-dependent evaporation rate, which is a function of temperature *T*_*t*_, humidity *H*_*t*_ and a physical constant *C*:

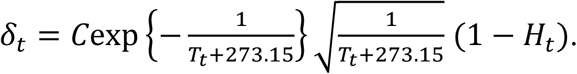

Then the expected duration of habitat at time *t i*s:

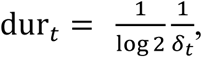

and we may approximate Habitat(*t*) as HSI(*m*) for a month *m* of length |*m*|:

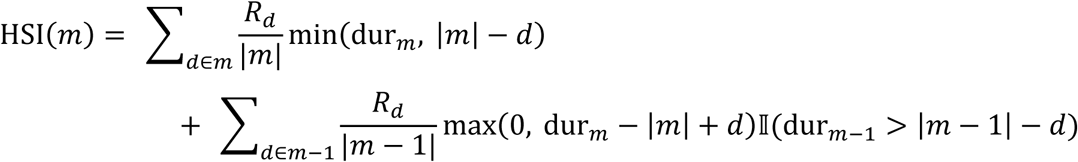

where 𝕀 denotes an indicator variable equal to 1 when the condition is true and 0 otherwise. We augmented this transient larval habitat suitability with a ‘permanent larval habitat’ layer derived from Tasselled Cap Wetness^60,66^ observations adjusted for local rainfall.

To visually examine the empirical signal of these two climatic suitability indices, we binned *Pf*PR observations into deciles by TSI and HSI (Extended Data Fig. 7). Subsetting observations to those in rural settings, and the further to areas of low ITN coverage, the variation in *Pf*PR associated with changing TSI and HIS becomes more pronounced.

### 3. Preparation of historical geotemporal housing, intervention, and contextual data

#### Malaria control

The scale-up of malaria control is responsible for most of the decline in burden in sub-Saharan Africa since 2000^36^. Geotemporal estimates of historic insecticidal bednet (ITN), indoor residual spray (IRS), seasonal malaria chemoprevention (SMC) and antimalarial treatment coverage were obtained from the Malaria Atlas Project. To account for the nonlinearity in the ITN-*Pf*PR_2-10_ relationship, ITN coverage was transformed using a pre-defined parametric interaction with estimated pre-intervention-era parasite rate^36^.

This study did not seek to model possible future trends in intervention coverage. Instead, we defined a baseline intervention coverage reflecting present-day levels, and this was used for all future scenarios (with or without disruption due to extreme weather events). To account for the campaign-based nature of ITN distribution we took a four-year average across 2019-2022 as baseline. For treatment coverage, delivered horizontally, 2022 was taken as baseline for future projections.

#### Relative abundance of vector species

Malaria ecology varies by mosquito species, with the relationship between the environment and transmission expected to vary between settings with different dominant vectors, even after controlling for interventions and socioeconomic factors. To account for this we fit species-specific terms in the model, with predictions based on relative abundance of the three dominant vectors in sub-Saharan Africa: *Anopheles gambiae* (s.s. and *coluzzi*)*, Anopheles funestus,* and *Anopheles arabiensis*. Estimates of relative abundance of each species were obtained from Sinka *et al*^49^, providing for each 5×5 km grid cell a three-way weighting of malaria vector species. We omit explicit modelling of the invasive *Anopheles stephensi* vector but acknowledge this mosquito represents a substantial emerging threat.

#### Socioeconomics

Improved housing has been shown to reduce risk of malaria infection^67^ and is a key mechanism by which socioeconomic development impacts malaria independently of direct malaria investment. We updated existing estimates of the prevalence of improved housing^38^ using data on characteristics of 1,083,386 households across Africa^68^ and predictors including gridded data on population density^56^, GDP^69^, land cover^70^, and travel time^71^. To account for the lifespan of buildings, a monotonicity constraint (increasing) was applied. We did not seek to model future trends in improved housing, and 2022 was used as the present-day baseline for projections.

### 4. Preparation of historical malaria response data

49,994 geo-located observations of *Plasmodium falciparum* parasite rate (*Pf*PR) were collated from DHS Program and other nationally representative surveys^68^ and systematic literature review^51^, representing 2.54 million individuals tested between 1995 and 2021 across 41 African countries. These data were standardised for age (to 2-10 years) and diagnostic type (to light microscopy), consistent with established approaches to modelling these data^72^.

### 5. Characterising historical climate, housing, intervention and other contextual effects on malaria

We used stacked generalisation^73^ to regress the predictor variables onto *Pf*PR observations. This method – which ensembles out-of-sample predictions from multiple ‘level 0’ models as predictors in a final geostatistical generaliser – has been shown to outperform standard model-based geostatistics when applied to *Pf*PR data^73^.

Three ‘level-0’ models were used in this analysis:

1. Linear model: for observed *Pf*PR *y*,

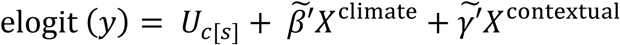 where *U*_*c*_ was a country specific intercept, 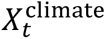 was a matrix with columns (i) standardised asinh-transformed HSI, cumulative across two- and three-month lags; (ii) standardised TSI, at one month lag, (iii) their multiplicative interaction; (iv) and (v) *An. arabiensis* relative abundance weighted versions of (i) and (ii); and finally (vi), (vii) *An. funestus* relative abundance weighted versions of (i) and (ii) [i.e. *An. gambiae* (s.s. and *coluzzi*) was taken as our reference species for terms (i) and (ii)]. 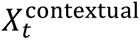 was a five-column matrix consisting of prevalence of improved housing; the ITN interaction term; IRS coverage; access to effective treatment with an antimalarial; and SMC coverage. elogit denotes the empirical logit.
2. Generalised Additive Model: Model formula as for meta-model (1), with *An. funestus* as reference species, and the linear terms for *An. gambiae* relative abundance weighted HSI, housing, IRS and SMC replaced with penalised cubic splines.
3. Generalised Boosted Regression: Model formula as for meta-model (1), with interaction term removed and replaced with species-specific terms. Monotonicity constraints were placed on the model terms to prevent spurious results. 1000 trees were fit, with an interaction depth of four.

These three models were fit to the *Pf*PR_2-10_ dataset described above, avoiding overfitting by using five-fold cross-validation to generate out-of-sample predictions to be used as fixed effects when training the level-1 generaliser model. For tractability, we modified the stacked generalisation approach by, for each level-0 model, generating these hold-out predictions with interventions set to zero. That is, each of the level-0 models provided an (out-of-sample) predictor representing estimated risk in the absence of interventions.

Each intervention class was then included in the geostatistical primary model as a fixed effect. For location *ii* and time *t* the generaliser model can be expressed, for *Pf*PR *y*_{*s*,*t*}_, as:

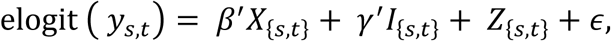

where *X*_{*s*,*t*}_ are the out-of-sample zero-intervention predictions forming the level-0 stack, *I*_{*s*,*t*}_ is the vector of malaria control coverage (interacted with pre-intervention-era *Pf*PR, in the case of ITNs) at pixel *ii* at time *t*, *Z* is spatio-temporal random field with separable Matérn-5/2 spatial and AR(1) temporal kernel, and *∈* is uncorrelated measurement error. The meta-learner slopes *β*^′^ were non-negative, a sufficient condition for stacked generalisation^73^. Following previous approaches to modelling *Pf*PR, we used a Gaussian likelihood and empirical logit transform to ensure well-specified posterior distributions^74^. The geostatistical model was fit in R 4.2.0 using INLA v22.12.16^75,76^. Fitted parameters for both the level-0 models and geostatistical generaliser model are given in the Supplementary Annex.

#### Validation

In-sample observed versus fitted correlation was 0.86 and mean absolute error was 8.5% at cluster and 1.1% at DHS survey aggregate. Verification of out-of-sample predictive ability using five-fold cross-validation yielded out-of-sample correlation of 0.83, mean absolute error of 9.5% (2.5% DHS survey out-of-sample aggregate).

### 6. Generation of future scenarios of extreme weather events

#### Scenario-based projections of flood events

Random Forest models were developed to predict flood occurrence, extent and duration. The training set consisted of historic flood events extracted at 230m spatial resolution from Floodbase^52^, aggregated to Pfafstetter Level 4 basin and over-sampled to reduce bias arising from data imbalance. A binary classifier was trained on historic Level 4 basin flood occurrence using predictors including rainfall and Atlantic and Indian Oceans sea-surface temperatures (see Supplementary Annex Table 6 for a full description of 27 predictors).

Flood extents (in km^2^) and durations were modelled probabilistically with similar predictors (see Supplementary Annex Tables 7, 8 for full description).

The performance of the three flood models was assessed using a 30% test set. The frequency model correctly classified 82% of occurrences and 85% of non-occurrences (AUC = 0.84), whilst the duration and extent models had *R*^2^ of 0.83 and 0.81, respectively.

We calculated GCM-specific future projections of flood occurrence, duration and extent by L4 basin for 2024-2049. For each GCM, the simulated period 2024-2026 was resampled to generate future counterfactual scenarios representing present-day climate flood frequency and extents. To downscale the predicted L4 basin-level extents, high-resolution occurrence data of historic floods^52^ were combined with a hydrological model^77^ to obtain, for each grid cell, flood propensity scores. For each flood event, grid-cells were then flooded from highest to lowest flood propensity until the predicted extent was reached. Projections were *ceteris paribus*, with land use variables held constant.

#### Scenario-based projections of cyclones

Historical Indian Ocean cyclone data (track morphology, start and end date, wind speeds) were obtained from IBTrACS^53^. Storms in the IBTrACS database coming within 50 km of the coast of Africa were included, yielding a training set of 192 tropical depressions, storms, and cyclones since 1980. Future scenarios of cyclone genesis and trajectories were generated using the Imperial College Storm Model (IRIS) dataset^54^, a 10,000-year synthetic dataset of statistical characteristics of cyclones, adjusted with climate data from the downscaled and bias-corrected CMIP6 models. The spatial probability of Lifetime Maximum Intensity (LMI) was modelled using coordinates of historic cyclone LMIs and observance of Poisson’s law. From these LMIs, synthetic tracks were generated by perturbing historical tracks using forecast cone uncertainty.

The maximum sustained wind speeds at LMI were calculated using climate data and thermodynamic constraints, including potential intensity (PI). After LMI, the model simulated intensity decay separately for ocean and land. Track steering and wind speed calculations used decay rates based on observed data and projected climate variables. Cyclone size was calculated dynamically, starting from LMI using a radial wind profile evolving along the track, capturing intensity-dependent size changes. Minimum pressure during the decay phase was modelled using a unified pressure-wind relationship, influenced by storm size and latitude. These components together generated synthetic cyclone datasets which replicated key physical and statistical characteristics of observed cyclones.

Only Category 1 or greater cyclones making landfall in Africa were included in our final scenarios. Cyclones were assumed to begin at LMI, and tracks were terminated when modelled wind speed fell below tropical storm threshold (63 km/h). A counterfactual future scenario of cyclones reflecting present-day climate conditions was generated by resampling past cyclones, with Poisson rates of each category given by their frequency since 1980. Impact footprints were calculated based on R_18_, the radius of damaging winds.

### 7. Parameterising impact of extreme weather events on housing and interventions

Extreme weather events were modelled as disrupting access to three key suppressants of malaria transmission: (i) improved housing, (ii) indoor vector control tools, and (iii) access to antimalarial treatment. The magnitude and duration of disruption was parameterised for each event class and severity based on a mixed methods approach: a literature review extracted 22 studies from the peer-reviewed and grey literature documenting the impact of extreme weather events (Extended Data Table 2). This process was augmented with 35 expert interviews. Parameters derived from this exercise were used to define recovery curves to be applied within footprints of extreme events, shown schematically in Extended Data Fig. 2, with parameters described in Extended Data Table 3. Acute and persisting impacts were differentiated, with the latter parameterised sigmoidally by time to 50% and 99% recovery. Uncertainty in the magnitude of disruption was then represented using a 50-150% scaling around the consensus central values.

### 8. Projecting impact of extreme weather events on housing and interventions

#### Disruptions to healthcare accessibility

Present-day access to effective treatment (EFT) for clinical malaria at location *ii* was estimated using a composite of three surfaces generated by the Malaria Atlas Project^51^: probability of care-seeking *c_s_* ; use of antimalarial drugs 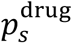; and drug- and location-specific therapeutic efficacy 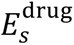:

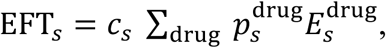

where ‘drug’ is one of Artemisinin Combination Therapy (ACT, the first-line treatment for *falciparum* malaria in Africa), Amodiaquine, Sulfadoxine-Pyrimethamine, Chloroquine or quinine.

Using a database of health facility geolocations^78^, a transport network model for Africa^79^ and a least-cost-path journey time algorithm^71^, we determined present-day (disruption free) travel time to health facilities for each grid cell. A bi-exponential relationship between these travel times 𝒯*s* and 104,516 geo-located observations of propensity to seek care for fever was fitted, resulting in the function:

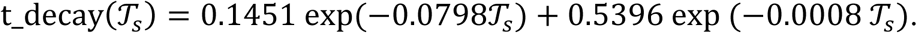

Health facilities located within the footprint of extreme weather extents were considered non-functional during acute impacts, with the proportion of facilities in each 5×5 km grid cell remaining non-functional in the post-acute period sampled from a binomial distribution, with probability of closure given by intersecting the appropriate recovery curve with a time-since-last-event surface. Given these dynamic functional facility geolocations, we recalculated travel time to health facilities for each scenario, by month, to 2050. In addition to facility closures, damage to road infrastructure was parameterised as travel time penalties derived from the recovery curves and time-since-event surfaces overlaid on the OpenStreetMap road network^79^. Off-road travel time was recalculated by perturbing a friction surface^78^ in the same way. The result was scenario-specific travel-time to healthcare 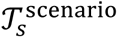. We then calculated care-seeking penalties relative to undisrupted conditions, so that at location *i* and time *t*:

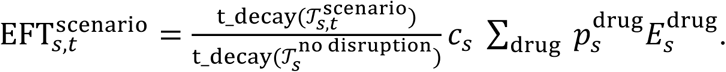

Antimalarial efficacy, proportional usage of different antimalarials, and secular variation in care-seeking behaviour were held constant, as was the undisrupted transport network.

#### Disruptions to ITN coverage and access to improved housing

ITN campaigns were simulated every three years on January 1^st^, commencing in 2025. As we aimed to model unmitigated impacts of extreme weather events, disrupted ITN coverage did not return to normal until the next simulated campaign. ITN access was assumed to be lost if access to housing was acutely disrupted. Access to improved housing was directly perturbed using the derived recovery curve.

### 9. Projecting ecological and disruptive effects of climate change

A set of scenarios was defined to derive the (a) ecological; (b) disruptive; and (c) combined impacts of climate change by the 2040s. These are described in Extended Data Table 1.

To model ecologically-driven impacts of climate change we generated estimates of monthly *Pf*PR_2-10_ from 2019 to 2049 for each of the 14 ensemble members. A climate-change free scenario (E_0_) was defined as the ensemble mean of median *Pf*PR_2-10_, 2019-2022, and corresponding scenario of future changes in ecological suitability (E_1_) as ensemble mean of median *Pf*PR_2-10_, 2040-2049. The ecological impact of climate change was thus estimated by the difference E_1_-E_0_ (Fig 1a).

We derived counterfactual and SSP 2-4.5 extreme weather event scenarios for a tractable subset of three GCMs: ACCESS-CM2, EC-Earth3-Veg-LR and MPI-ESM1-2-LR. For each ensemble member, two time-series of spatial *Pf*PR_2-10_ projections were calculated. Scenario D_0_ was defined as the (three-GCM) ensemble mean of median *Pf*PR_2-10_, 2040-2049, generated with present-day TSI and HSI as in E_0_ and with extreme events simulated as occurring at present-day frequency. Still holding TSI and HSI at present-day levels, SSP 2-4.5 projections of changing extreme weather event frequency were imposed to derive scenario D_1_, so that the difference D_1_-D_0_ isolated the impact of disruptive events due to climate change (Fig 2a, Extended Data Table 1).

Ecological and disruptive impacts were combined by defining C_0_ equal to D_0_ (i.e. present-day climate suitability and extreme event frequency). SSP 2-4.5 projections of changing climate suitability indices (as in E_1_) and changing extreme weather frequency (as in D_1_) were combined to generate C_1_, the (three-GCM) ensemble mean of median *Pf*PR_2-10_, 2040-2049. The combined ecological and disruptive impacts were then calculated as C_1_-C_0_ (Fig 2b).

Projected *Pf*PR_2-10_ for each scenario was converted to clinical case incidence using an established natural history model^55^. Gridded population projections consistent with SSP 2-4.5 were used to generate estimates of absolute cases^56^. Projected time-series of clinical cases and mortality were scaled to align with 2022 official World Health Organisation estimates, as reported in World Malaria Report 2023^57^.

## Data Availability

The downscaled and bias-corrected CMIP6 climate projections used in this analysis are available from: https://registry.opendata.aws/nex-gddp-cmip6/. Citations to other supporting datasets (on historical climate variables, flood and hydrological modelling, cyclone modelling, topology, population density, remotely sensed landcover classifications, socioeconomic indicators, malaria infection prevalence and control coverage) are provided in the manuscript or in full in Supplementary Annex Table 1.

---

**Extended Data Figure 1.**
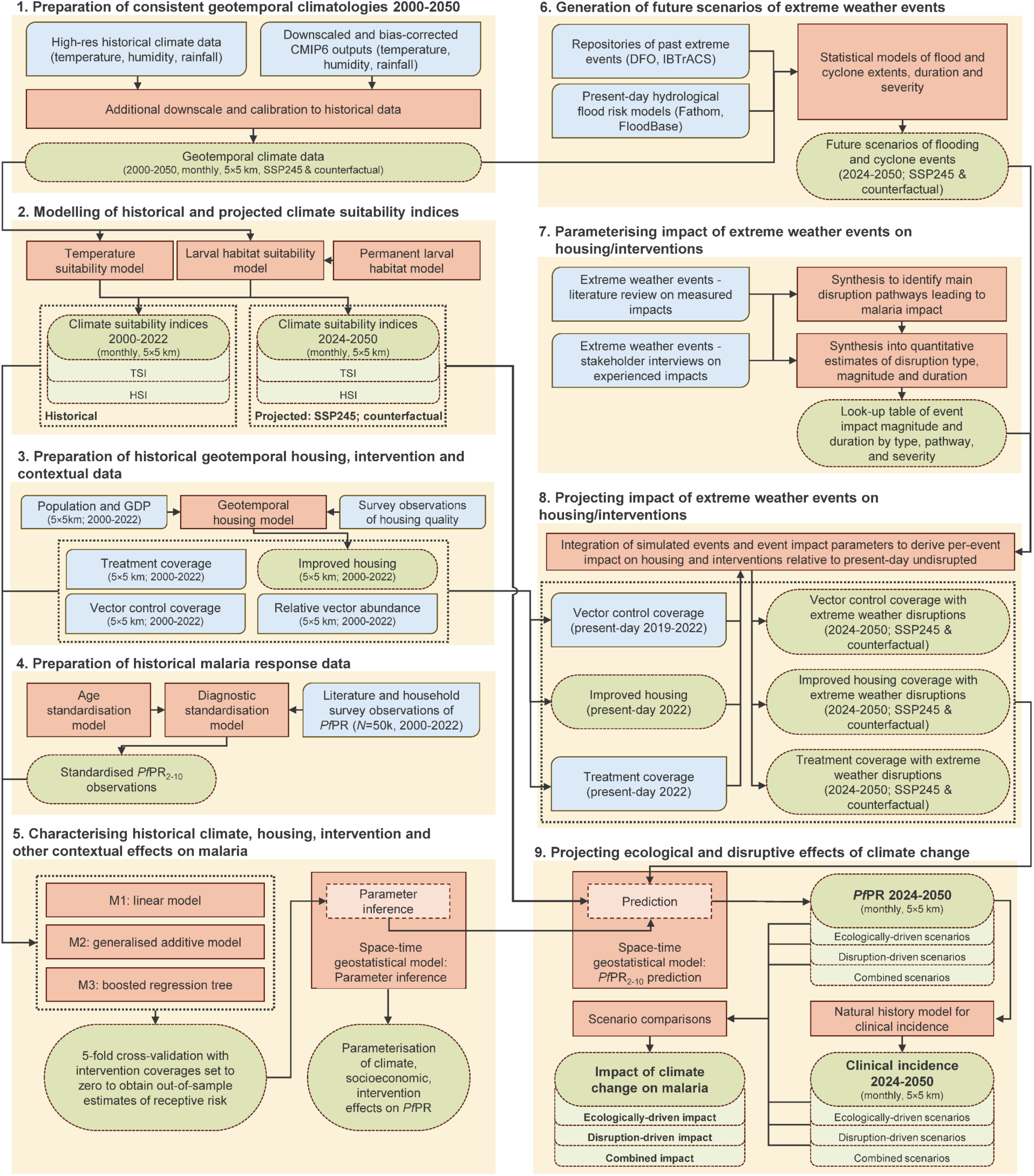
Schematic overview of input data, model components, and outputs. Each analysis stage is described in the Extended Methods, with additional detail in the Supplementary Annex. Blue boxes are input data; orange boxes are modelling or computational steps; green rods are interim or final outputs. DFO = Dartmouth Flood Observatory; IBTrACS = International Best Track Archive for Climate Stewardship; TSI = Temperature Suitability Index; HSI = Habitat Suitability Index; *Pf*PR = *Plasmodium falciparum* parasite rate.

**Extended Data Figure 2.**
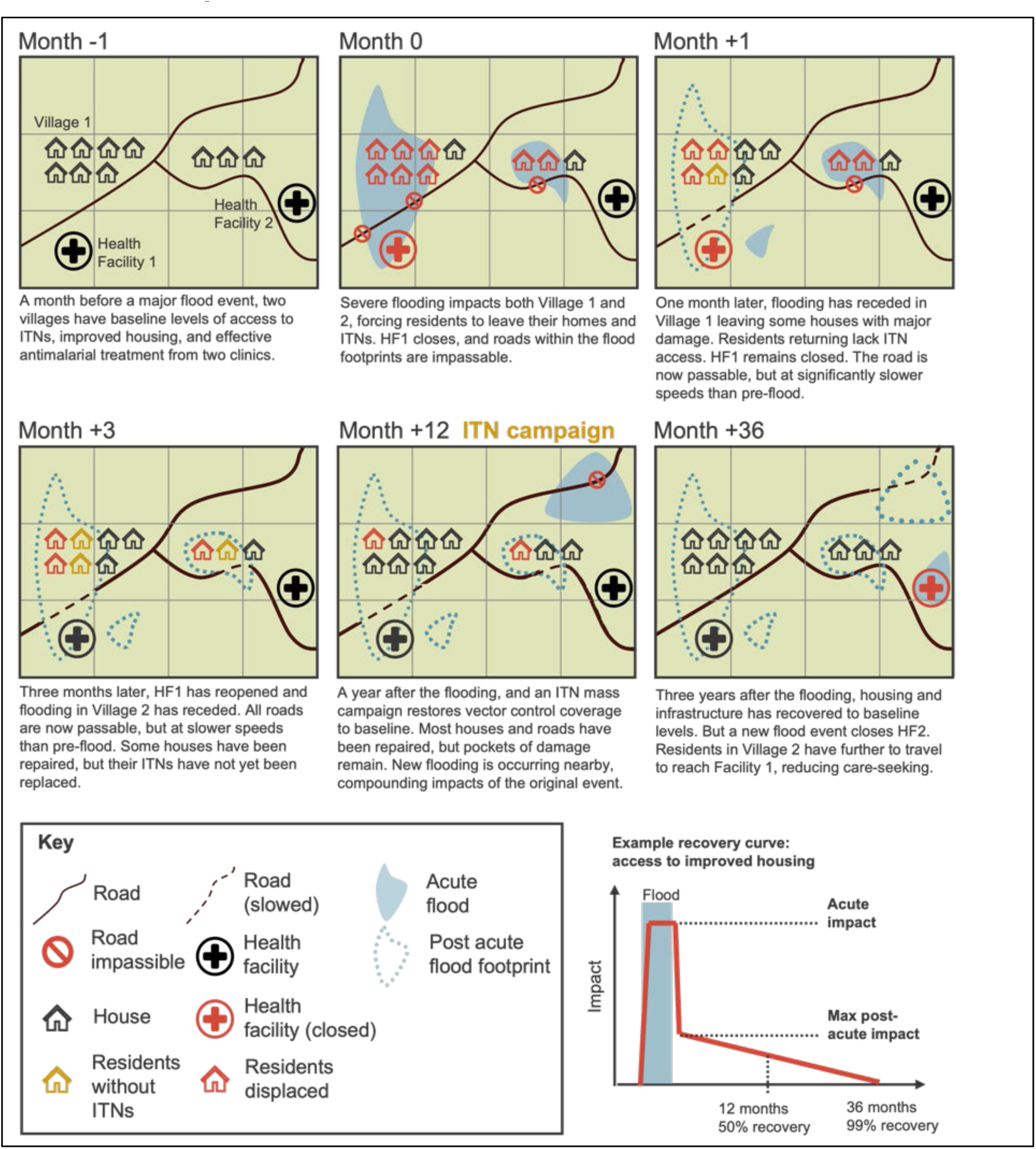
Schematic overview of extreme event impact methodology. Panels show the progression of modelled disruptive effects of flooding from pre-flood baseline (Month −1), through the acute flooding period (Months 0, 1) and into the recovery phase (Months 3 to 36). Considered disruptive effects were disrupted road infrastructure (prohibition signs for impassable roads, dashed lines for passable but damaged roads), closures of health facilities (shown in red), damaged housing (red houses) and loss of access to vector control (yellow houses). An example recovery curve is plotted, showing both acute impacts (during flooding shaded in blue) and long-term return to baseline.

**Extended Data Figure 3.**
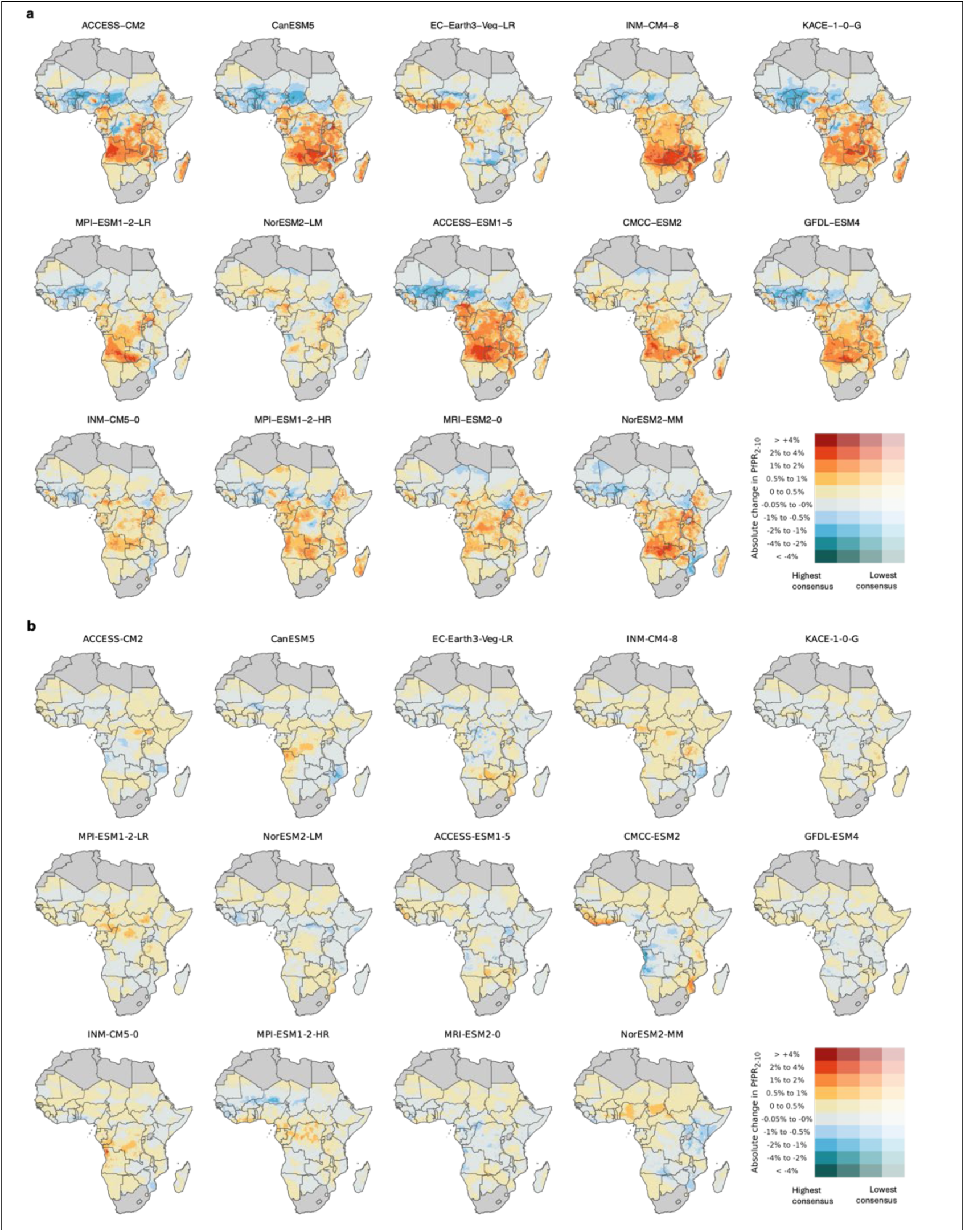
Projected climate suitability impacts, fourteen CMIP6 model ensemble. Each plot corresponds to a member of the (downscaled and bias-corrected) CMIP6 model ensemble used in the ecological impact analysis. Presented metrics are as in Figures 1 and 2: percentage point change in *Pf*PR_2-10_ median across 2040-2049, relative to a 2019-2022 baseline scenario, with **a** changing temperature suitability, with larval habitat suitability index, intervention coverage and socioeconomics held constant at contemporary levels in all scenarios, and **b** changing larval habitat suitability, with temperature suitability index, intervention coverage and socioeconomics held constant at contemporary levels in all scenarios

**Extended Data Figure 4.**
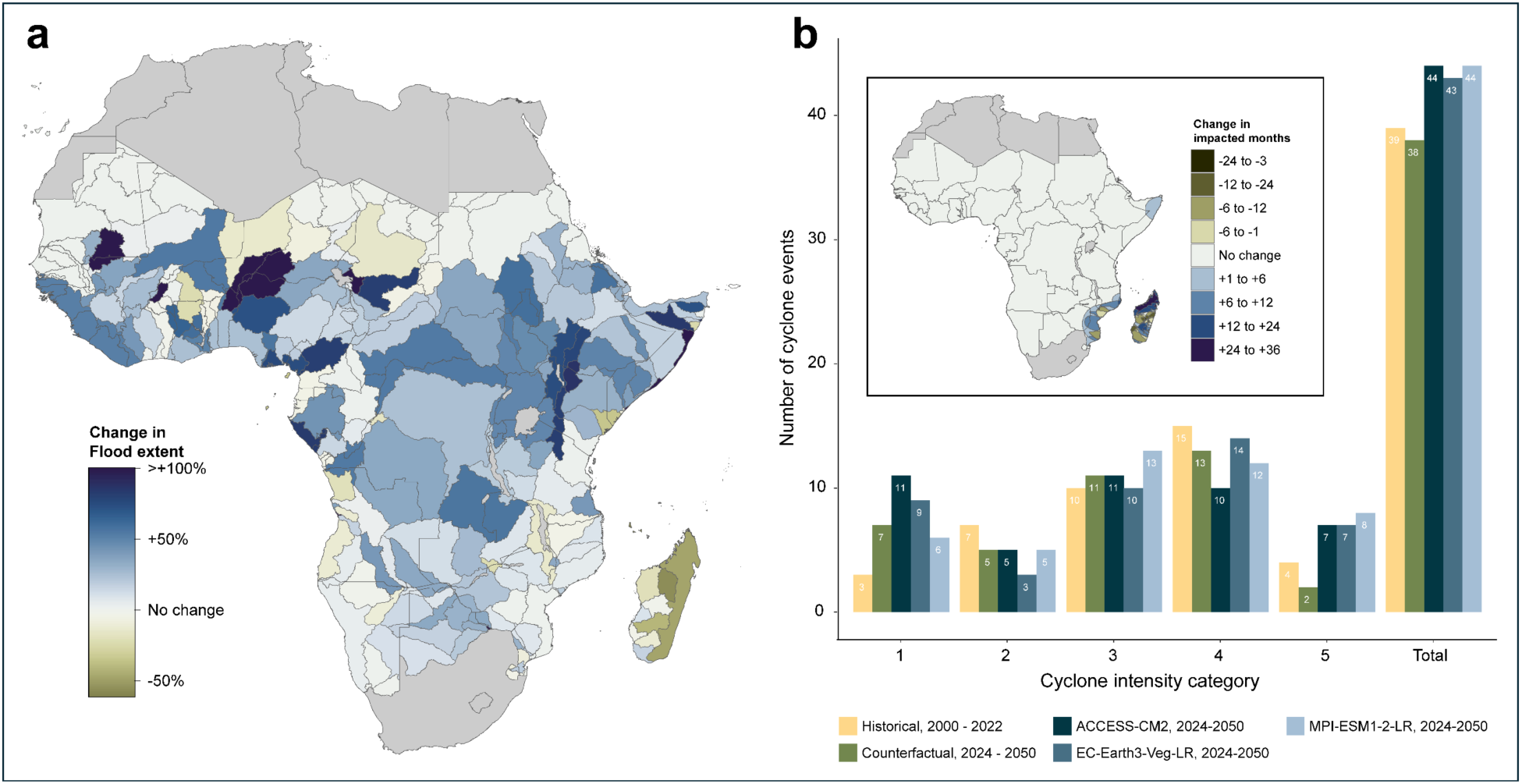
Projected change in flood and cyclone events under SSP245. **a** Mapped projected change in flood frequency in Africa in the 2040s. Metric shown is ACCESS-CM2, EC-Earth-Veg-LR and MPI-ESM1-2-LR ensemble mean percentage change in cumulative flood extent 2040-2049, where flood extent is defined as the proportion of each 5×5 km grid-cell flooded in each simulated month-year. Percentage change is relative to simulated baseline of flood extents given present-day statistics of flood occurrence, frequency, severity, and extent. **b** Historical and simulated cyclone event frequency by Category, 2000-2022 and future simulations under baseline, ACCESS-CM2, EC-Earth-Veg-LR and MPI-ESM1-2-LR projections. White annotations denote number of events in each model-category. Inset map shows change in number of number of months impacted by cyclones, summarised as ensemble mean delta (modelled scenario minus baseline scenario), cumulative across the 2040s.

**Extended Data Figure 5.**
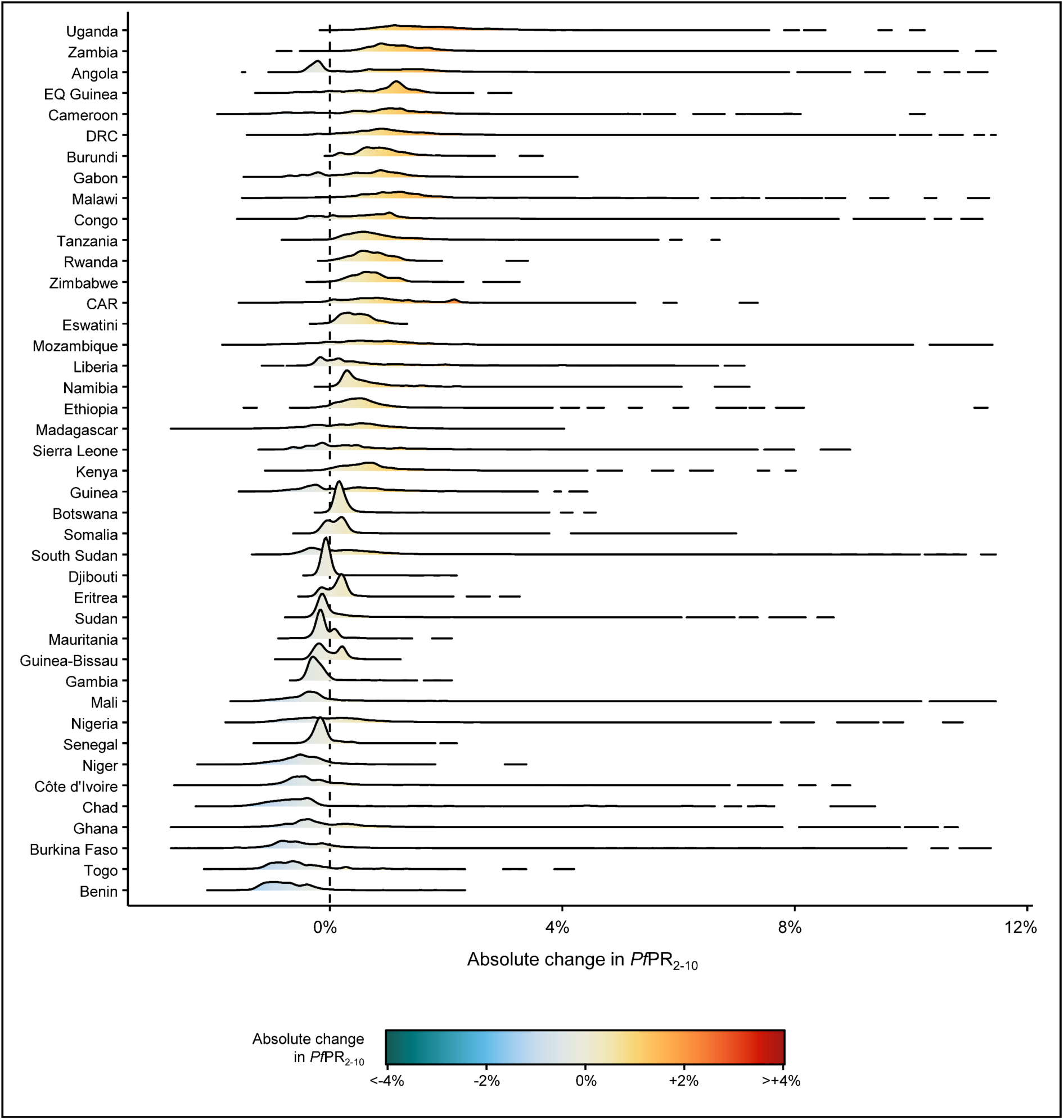
Distribution of ecological and disruptive climate change impacts on *Pf*PR_2-10_ by country. Each ridgeline shows, for a given country included in this analysis, weighted (by population and model ensemble consensus score) kernel density estimates of 5×5 km grid-cell absolute change in *Pf*PR_2-10_ in the 2040s due to combined ecological and disruptive impacts of climate change, SSP 2-4.5 (as mapped in Main Text Figure 2b). The y-axis is ordered by (descending) population- and consensus-weighted median change. The x-axis was clipped at 0.1/99.9% percentiles of overall change for visualisation purposes, whilst ridgelines with heights below 0.0001% of the maximum of their corresponding density estimate were censored. Densities were calculated with a joint bandwidth. EQ Guinea = Equatorial Guinea; DRC = Democratic Republic of the Congo; CAR = Central African Republic.

**Extended Data Figure 6.**
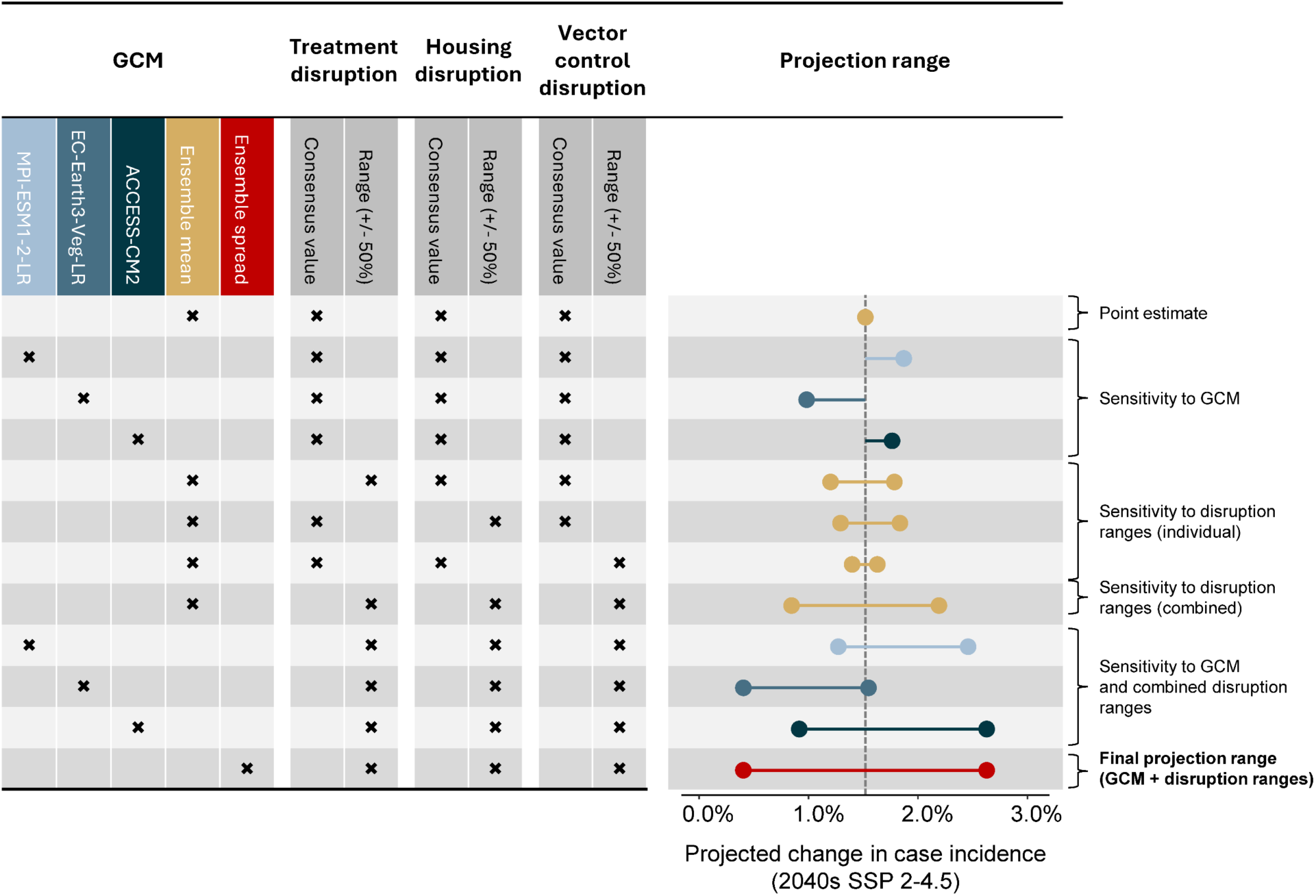
Sensitivity of projected climate change impacts to GCM model configuration and disruptive impact parameterisations. Horizontal bars show how estimates of climate change impact on case incidence rate in the 2040s vary under different model configurations. As specified in table on left-hand side, different model runs encompassed different GCM configurations (individual GCM choices, ensemble mean, ensemble spread), and reflected the uncertainty ranges (spanning 50% to 150% of the central consensus value) imposed around the parameters representing disruption to antimalarial treatment, housing and vector control. The top-most and bottom-most rows reflect, respectively, the configurations used for the point estimates and projection ranges presented in this study.

**Extended Data Figure 7.**
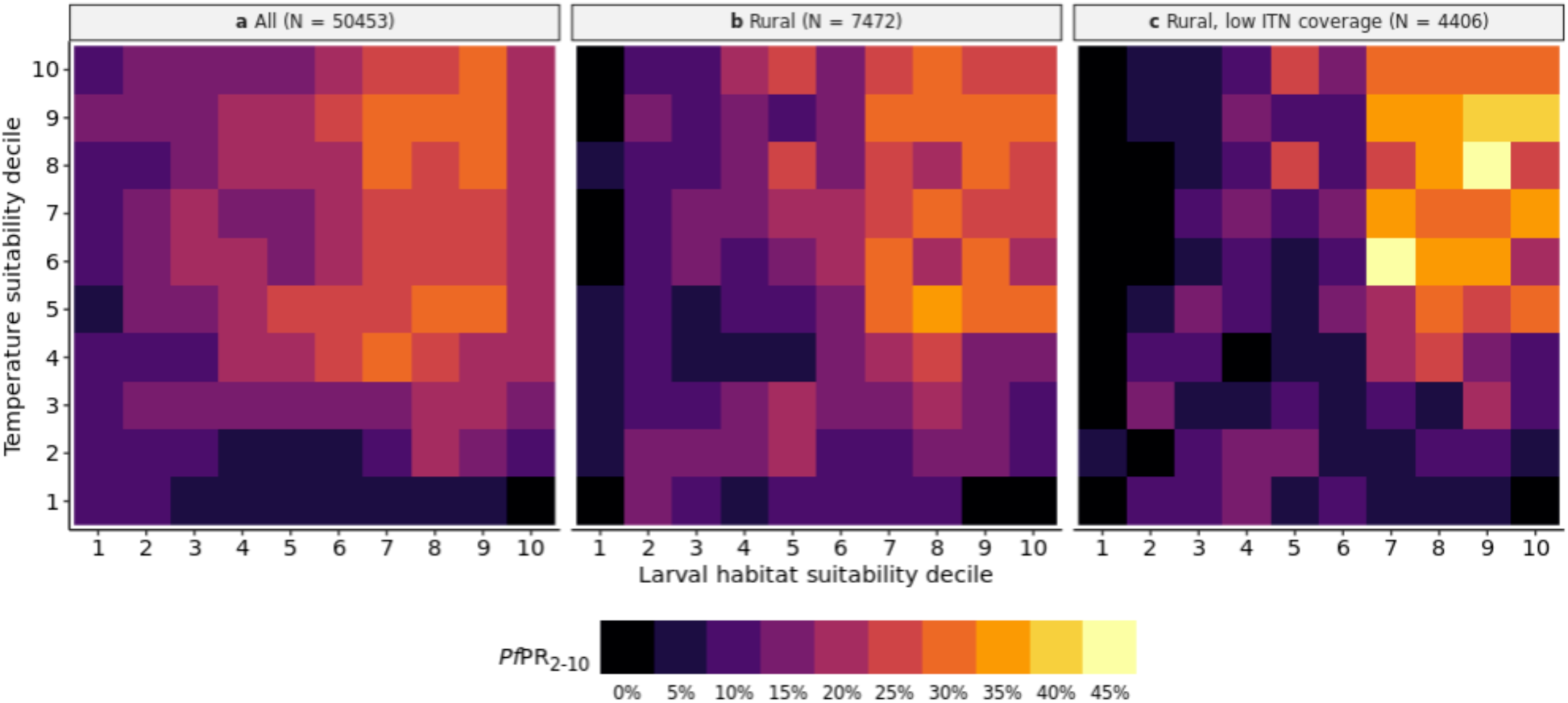
Observations of *Pf*PR_2-10_ by modelled climate suitability. Cross-sectional observations of *Pf*PR_2-10_ collected since 1995, binned into bivariate deciles of contemporaneous modelled larval habitat suitability (*x*-axis) and temperature suitability (*y*-axis). Higher deciles correspond to higher climate suitability. Panels correspond to **a** all geo-located *Pf*PR_2-10_ observations (*N*=50,453), **b** the dataset subset to rural locations (defined as GHSL built-up surface percentage <10% in the observed 5×5 km grid-cell, *N* = 7,472), and **c** a further subset of rural locations with low ITN coverage (rural defined as in panel b, additionally with estimated effective ITN coverage <35%, *N* = 4,406). *Pf*PR observations were standardised for age and diagnostic type but were otherwise unadjusted.

**Extended Data Table 1.** Summary of model configurations for evaluation of ecologically-driven, disruption-driven, and combined impacts of climate change on malaria. Each evaluation comprised a counterfactual scenario (E_0_, D_0_, C_0_) reflecting 2024-2050 without future climate change, and a comparator scenario (E_1_, D_1_, C_1_) reflecting 2024-2050 under climate change as projected via SSP 2-4.5, with the impact of climate change quantified as the delta (e.g. E_1_ - E_0_).

|  | Ecologically-driven impacts of climate change |  | Disruption-driven impacts of climate change |  | Combined impacts of climate change |  |
| --- | --- | --- | --- | --- | --- | --- |
|  | E<sub>0</sub> | E<sub>1</sub> | D<sub>0</sub> | D<sub>1</sub> | C<sub>0</sub> | C<sub>1</sub> |
| **Climate suitability indices – without future climate change** |  |  |  |  |  |  |
| TSI: derived from contemporary baseline climate (2019-2022) | ● |  | ● | ● | ● |  |
| HSI: derived from contemporary baseline climate (2019-2022) | ● |  | ● | ● | ● |  |
| **Climate suitability indices – with future climate change (SSP2-45)** |  |  |  |  |  |  |
| TSI: derived from SSP2-45 |  | ● |  |  |  | ● |
| HSI: derived from SSP2-45 |  | ● |  |  |  | ● |
| **Housing and malaria interventions – no extreme weather disruption** |  |  |  |  |  |  |
| Vector control: population coverage held constant at 2019-2022 levels | ● | ● |  |  |  |  |
| Treatment: population coverage held constant at 2022 levels | ● | ● |  |  |  |  |
| Improved housing: population coverage held constant at 2022 levels | ● | ● |  |  |  |  |
| **Housing and malaria interventions – disrupted by extreme weather without future climate change** |  |  |  |  |  |  |
| Vector control: 2019-2022 coverage with contemporary extreme events imposed |  |  | ● |  | ● |  |
| Treatment: 2022 coverage with contemporary extreme events imposed |  |  | ● |  | ● |  |
| Improved housing: 2022 coverage with contemporary extreme events imposed |  |  | ● |  | ● |  |
| **Housing and malaria interventions – disrupted by extreme weather with future climate change (SSP2-45)** |  |  |  |  |  |  |
| Vector control: 2019-2022 coverage with SSP2-45 extreme events imposed |  |  |  | ● |  | ● |
| Treatment: 2022 coverage with SSP2-45 extreme events imposed |  |  |  | ● |  | ● |
| Improved housing: 2022 coverage with SSP2-45 extreme events imposed |  |  |  | ● |  | ● |
| **Other contextual factors (considered static)** |  |  |  |  |  |  |
| Permanent larval habitat: derived from historic synoptic mean (2001-2022) | ● | ● | ● | ● | ● | ● |
| Relative vector species abundance: 1985-2010 levels | ● | ● | ● | ● | ● | ● |

**Extended Data Table 2.**
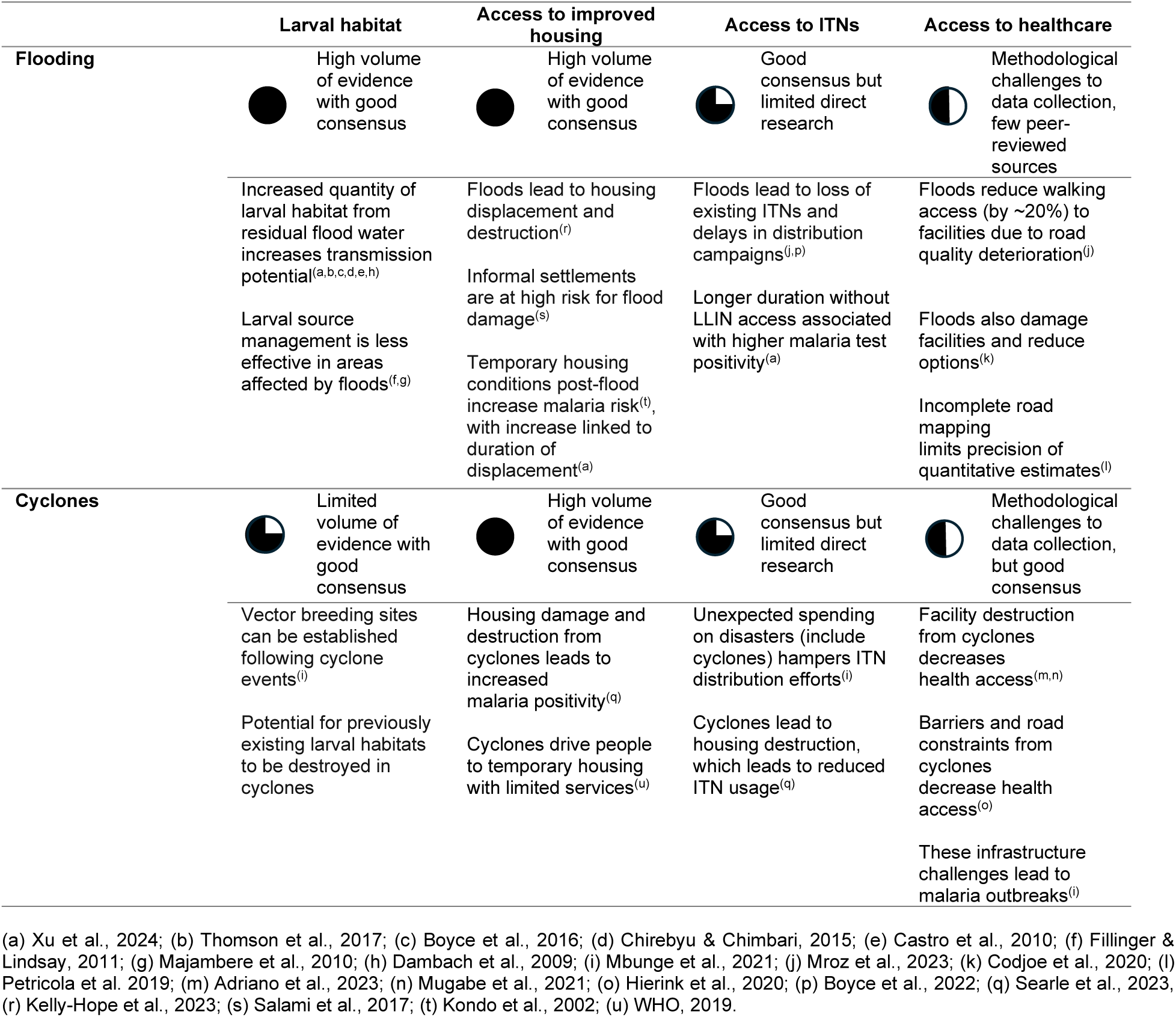
Summary of literature review on disruption to malaria control and access to healthcare following extreme weather events, including quality assessment. Studies are categorised by type of event and mechanism of disruption, with brief text summaries of findings presented. Harvey balls summarise qualitative assessment of volume, consensus, and robustness of evidence. Search terms and Harvey ball rubric are provided in the Supplementary Annex. Supporting references are provided in abbreviated form in the below footnotes, and in full in Supplementary Annex Section 7.1., Table 9.

**Extended Data Table 3.**
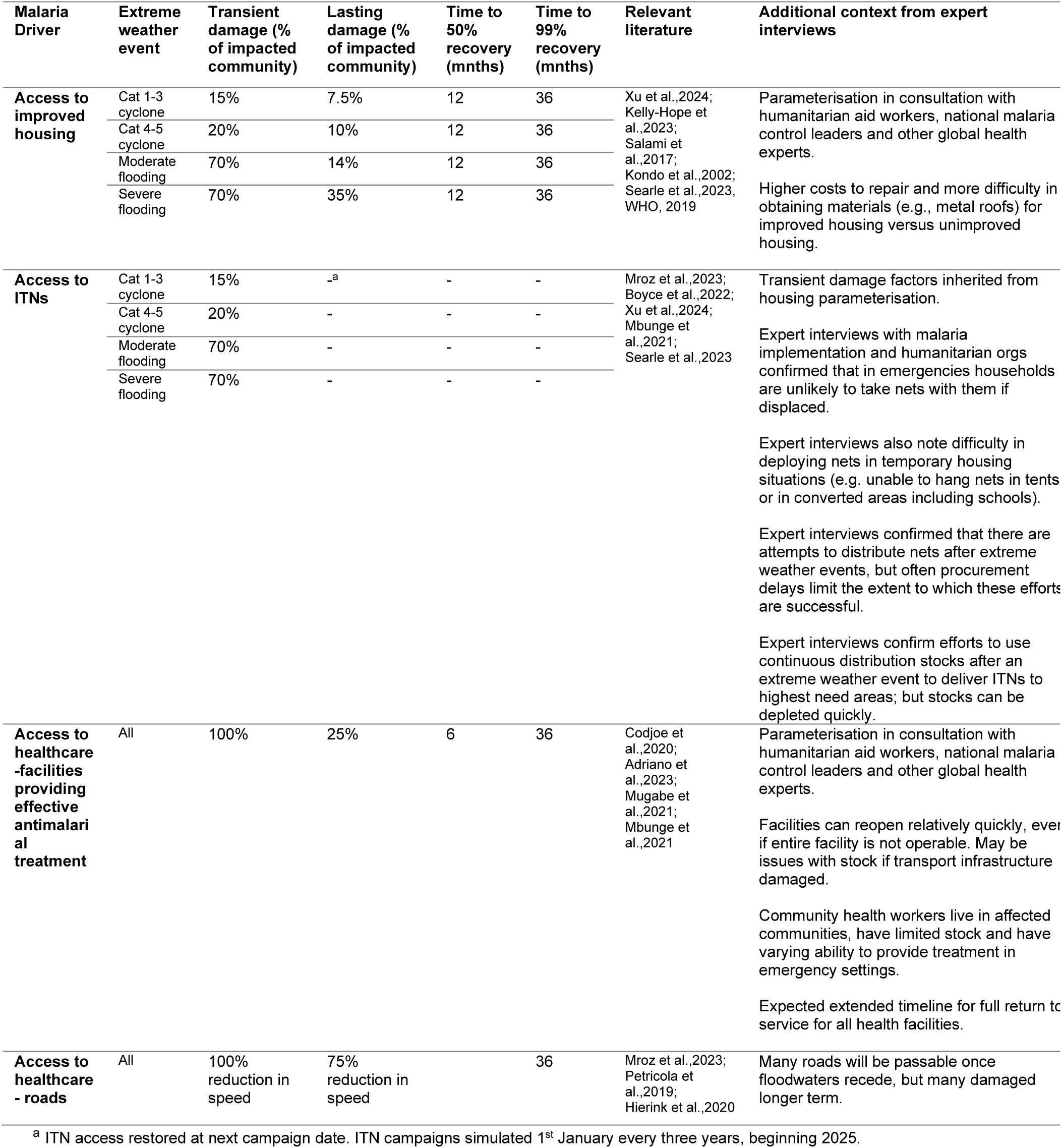
Parameterisation of impact of extreme weather events on malaria control. Damage and recovery parameters were chosen to represent approximate consensus across literature and expert stakeholders and were subsequently subject to a sensitivity analysis with broad ranges applied. Supporting references are provided in abbreviated form in the table, and in full in Supplementary Annex Section 7.1., Table 9.

